# Cultural values predict national COVID-19 death rates

**DOI:** 10.1101/2020.07.17.20156091

**Authors:** Damian J. Ruck, Joshua Borycz, R. Alexander Bentley

## Abstract

National responses to a pandemic require populations to comply through personal behaviors that occur in a cultural context. Here we show that aggregated cultural values of nations, derived from World Values Survey data, have been at least as important as top-down government actions in predicting the impact of COVID-19. Whereas trust in institutions predicts lower COVID-19 deaths per capita, secular-rationalism and cosmopolitanism each predict more deaths. The effects of these cultural values register more strongly than government efficiency. This suggests that open democracies may face greater challenges in limiting a pandemic, and that all nations should consider their cultural values as actionable parameters in their future preparations.

## Introduction

Combating the COVID-19 pandemic [1] in nations around the world has depended partly on efficient government response [2, 3, 4, 5, 6], and partly on the behaviors of individuals [7, 8, 9, 10]. Since culture is the context for behavior [10, 11, 12], effectiveness of government intervention on COVID-19 ought to reflect public trust in their institutions as well as other cultural values that vary substantially between countries [21, 14].

Here we estimate the observable effects of aggregated cultural attitudes in different countries on their COVID-19 fatality rates. Applying a two-stage factor analysis to World and European Values Survey data [19, 20] we previously derived cultural value factors—including secular-rationality (RAT), cosmopolitanism (COS) and institutional trust (INST)—in over one hundred countries [14, 15]. We entered these cultural values into a matrix of country-scale covariates, *X*, among the 87 counties for which we also have data for government efficiency, from an established index [26, 24], as well as logarithm of GDP per capita, population size, per cent urban population, and per cent of population aged 65 and over (see Materials and Methods).

We first explore the variance structure in the covariate matrix *X* by principal component analysis, to see how the principal components correlate with per-capita COVID-19 deaths. We then use multivariate regression to explain how the individual covariates predict the residual variance in COVID deaths in the different countries, 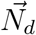:

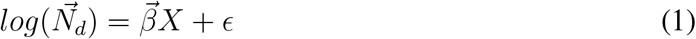

where the errors *ϵ* follow a negative binomial distribution and have a variance for a given mean, *µ*, of *µ*(1 + *µ/r*), where *r* is a dispersion parameter. The numbers of COVID deaths,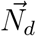 are daily-updated national counts [27]; we assume deaths to be a delayed proxy for true number of cases, but with less uncertainty [16, 25, 17].

Since COVID-19 deaths are count data that are highly dispersed, we model negative-binomial distributed errors, using the ‘glmmADMB’ package in R [23], rather than a more restrictive Poisson distribution. The regression determines the vector of country-specific coefficients, 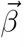, applied to the covariate matrix *X* (equation 1). While we consider the covariates as constants for the duration of the pandemic, the number of COVID-19 deaths grew exponentially so we fit regressions at one-day intervals from 5 to 55 days after the day when the first death occurred in each country. To ensure the results of these regressions are not sensitive to the 87-country sample, we find broadly the the same results using an ensemble of bootstrapped samples (see Supplementary Materials). We control for the demographic factors of population size, fraction urban population and fraction aged over 65 by including these as covariates in the regression.

## Results and Discussion

As Figure 1 shows, the initial spread of COVID-19 was positively correlated with cosmopolitanism, COS (Adjusted *r*^2^ = 0.229, *p* < 0.0001) and rational-secularism, RAT (Adj. *r*^2^ = 0.217, *p* < 0.0001). Deaths correlate negatively with institutional confidence, INST (Adj. *r*^2^ = 0.029, *p* = 0.066). There is surprisingly little correlation, however, with government efficiency (adj. *r*^2^ = 0.011, *p* = 0.168).

**Figure 1:**
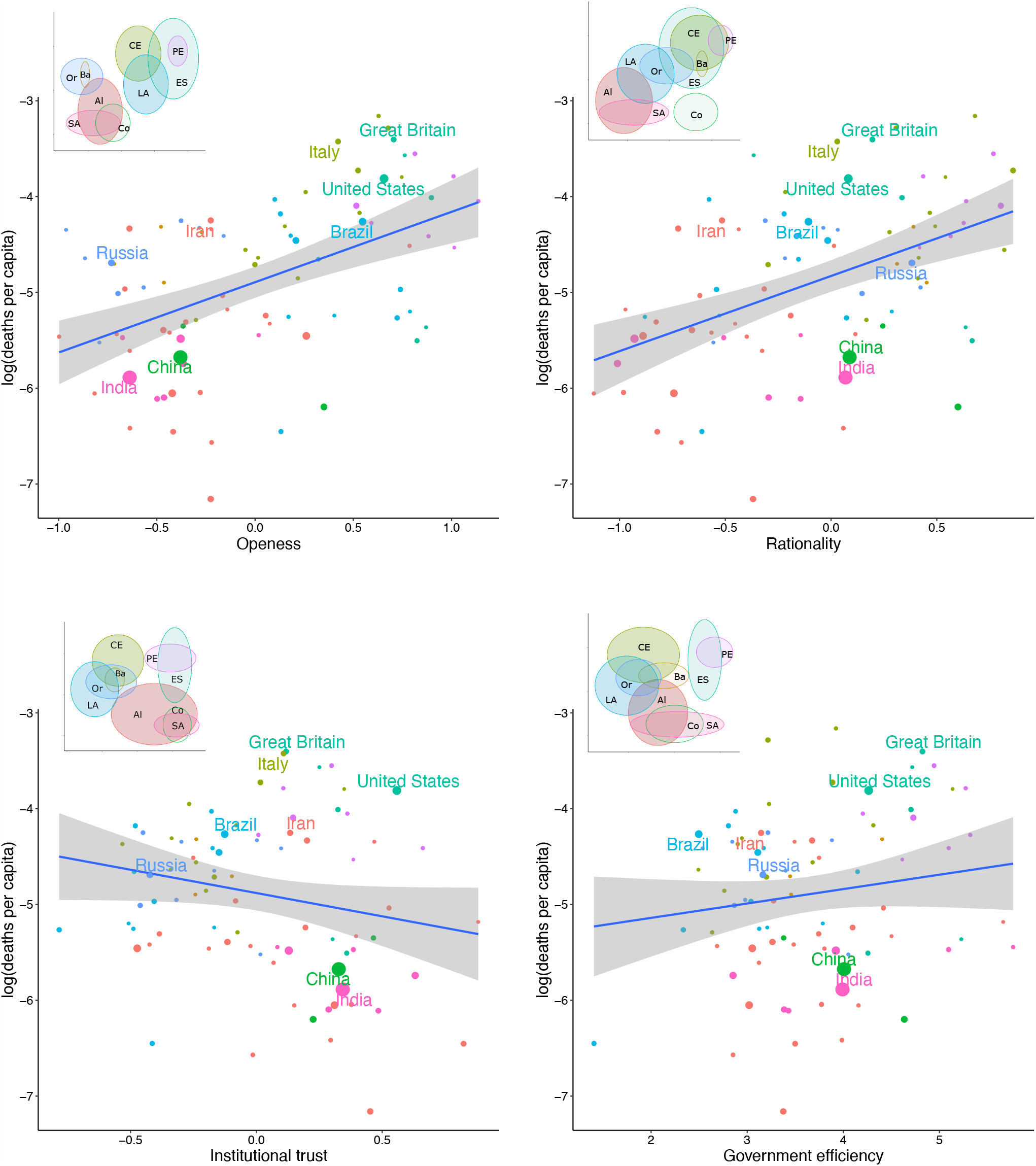
Bi-variate plots of Covid spread versus cultural values defined by [13]. Main plots show individual countries and LOESS correla1t0ion. Insets show 1 s.d. confidence ellipses by cultural region [15]: AI, African-Islamic; Ba, Baltic; CE, Catholic Europe; Co, Confucian; ES, English Speaking; LA, Latin America; PE, Protestant Europe; Or, Orthodox; SA, South Asia.

Principal component analysis (PCA) on all the variables appears to capture two main sources of variation in the first two PCs: socio-economic development and effect of institutions, respectively. Because the first component, PC 1, which accounts for 47% of the variance (Table S1), is positively loaded on all covariates (Table S2), we interpret it to be congruent with socio-economic development. The second component, PC2 (23% of the variance) is essentially documenting institutional confidence (INST) and government efficiency, which complement each other (Tables S1 and S2). Per-capita COVID-19 deaths correlate positively with PC1 and negatively with PC2 (Figure 3). This broadly suggests that socio-economic development has predicted more deaths while the effectiveness of institutions has reduced deaths.

In between these major components lie the cultural factors of cosmopolitanism and secular-rationalism (COS, RAT), which plot on a diagonal between PC1 and PC2 (Figure 2). These two principal components explain 70% of the total variance, and are loaded significantly on each of the seven covariates. Notably, PC4 (7.8% of the variance) is loaded almost entirely on COS (Supplementary Materials). In the Supplement (Figure S1) we show the effects on the PCA using additional, similar variables, such as Government Integrity, Judical Effectiveness, and Business Freedom (from the Heritage Foundation). These variables load primarily onto PC1, with Government Integrity having the biggest loading (Table S1). Figure 3 shows how COVID deaths correlate positively with PC 1 (Adj. *r*^2^ = 0.390, *p*< 0.0001) and negatively PC2 (Adj. *r*^2^ = 0.049, *p* = 0.024).

**Figure 2:**
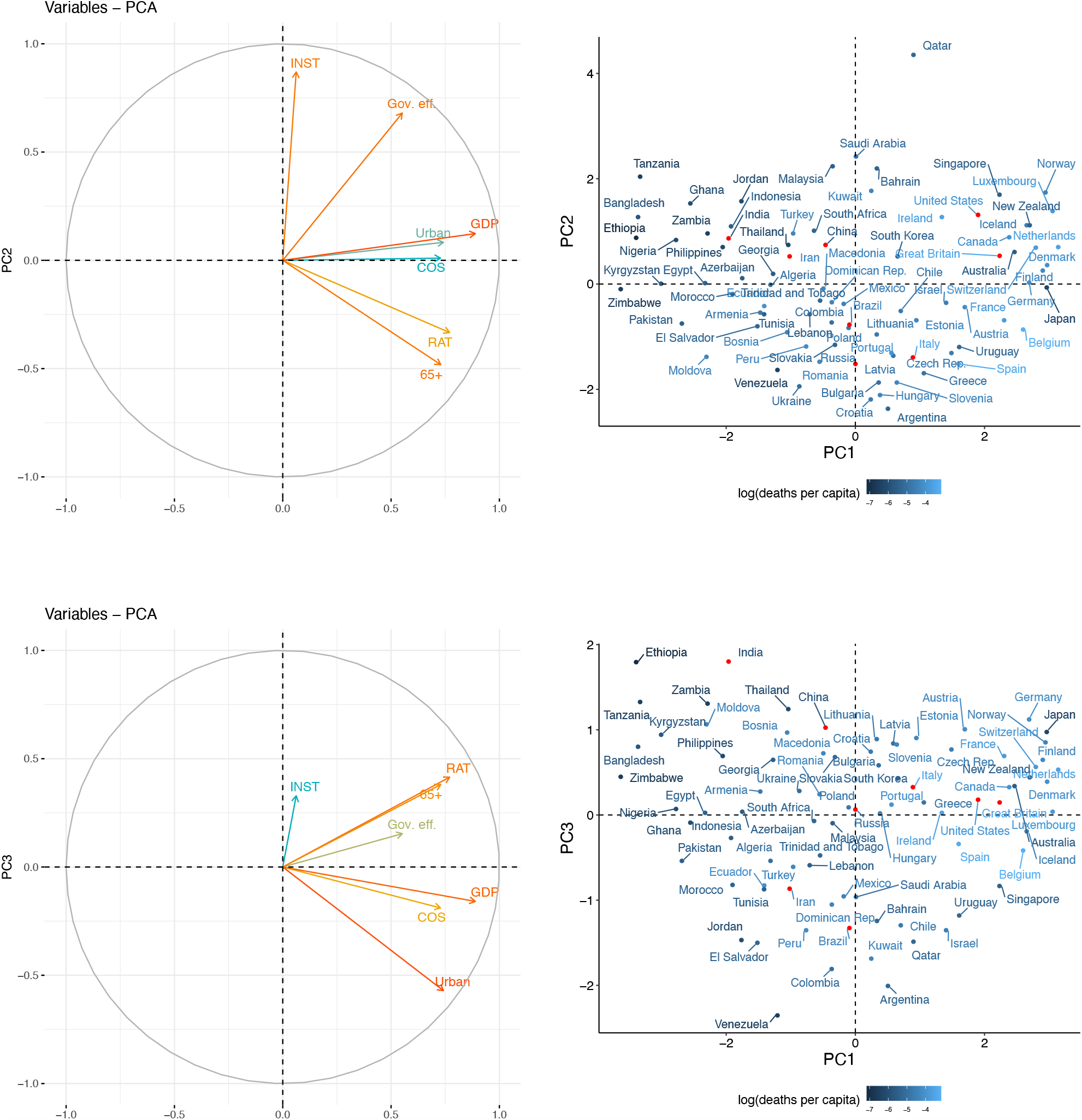
Principal components, PC1 vs. PC 2 on the top row and PC 1 vs. PC3 at bottom row. Plots on the left show how variables (see Table 1 for abbreviations) align with the PCs. Plots on right show PC scores for individual nations. Nations highlighted by red dots: Brazil, China, Great Britain, India, Italy, Iran, and U.S.

**Figure 3:**
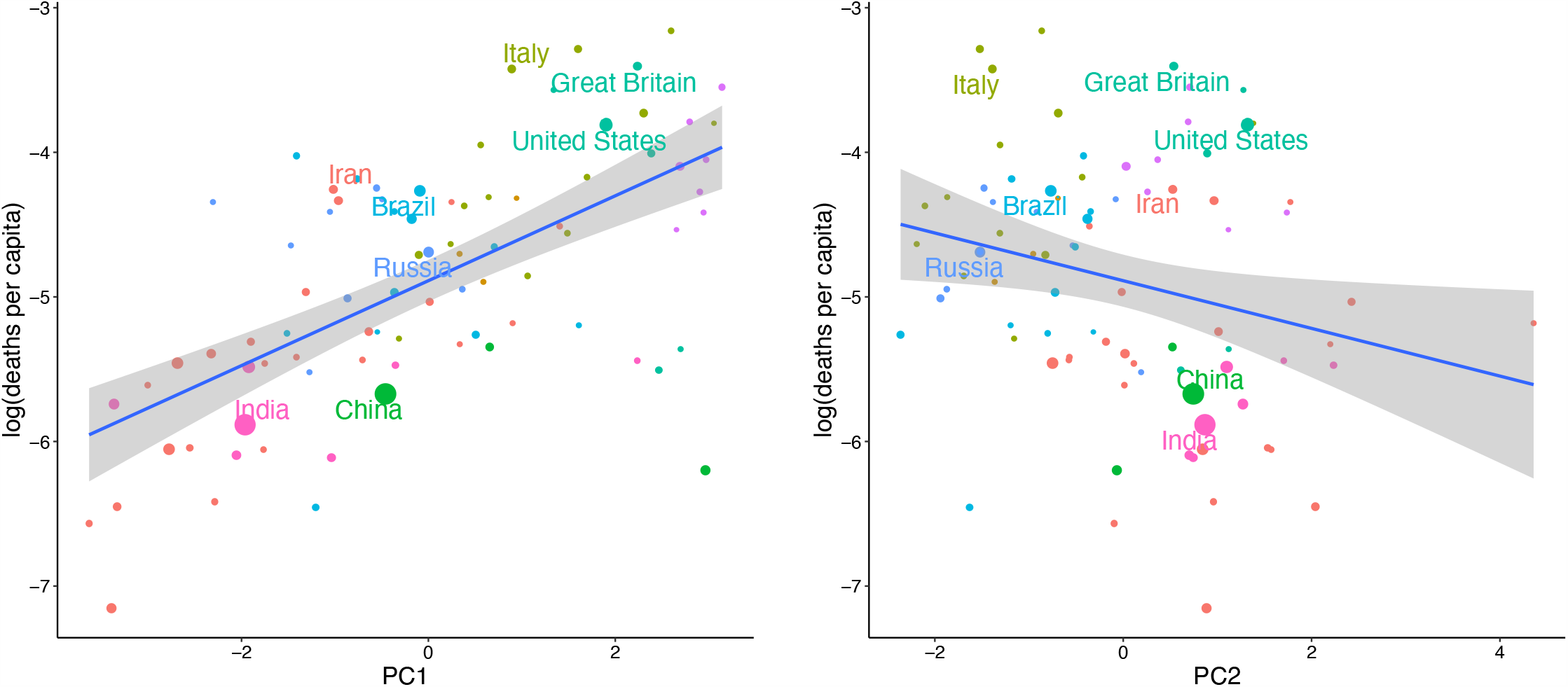
COVID-19 deaths vs **left** PC 1 and vs. **right** PC 2.

Having explored the principal components of variation, a multivariate regression helps disentangle the joint effects of cultural values, government efficiency and economic incentives. The effect of several covariates—COS, INST, GDP and government efficiency—increased between Day 10 and Day 50 (Table S4). The z-scores of these effects between 5 and 55 days after the outbreak (Figure S2) reveal a change in covariate effects over the first two months of the outbreak (since deaths are a delayed effect, the timeline may be offset by about two weeks). Both cosmopolitan openness (COS) and and GDP increased in significance, while secular-rationality became less important over the first two months (Figure S2). Institutional confidence (INST) remains near significance level (p = 0.05) for the entire two months, while government efficiency only became significant after the two months (Figure S2).

Table 1 summarizes the regression evidence that the aggregated cultural values of nations have been as important, if not more so, than government efficiency in COVID-19 death rates. At all stages during the COVID-19 outbreak, countries with strong trust in institutions (INST) had fewer deaths per capita. Trust in institutions (INST) presumably helps align health behaviours with scientific guidelines. Government efficiency, however, had only a weak effect, and not until the second month of the outbreak (Table 1). Including an interaction term in the multivariate regression did not add explanatory value, implying independent effects of government efficiency versus institutional confidence (Supplementary Materials). The independence may reflect the international nature of of medical evidence and advice for people to trust versus the local efficiency of national governments attempting specific orders and policies.

**Table 1:** Results of negative binomial regression. Predictors of COVID-19 deaths, 10 and 50 days after outbreak. In parentheses are heteroskedastic standard errors (for negative-binomial-distributed errors). Heteroskedastic adjusted significance: ^***^*p*< 0.001, ^**^*p*< 0.01, ^*^*p*<0.05, †p < 0.10

| Covariate (in matrix $X$) | Day 10 | Day 50 |
| --- | --- | --- |
| RAT | $-0.58 (0.31)^\dagger$ | $0.08 (0.43)$ |
| COS | $0.04 (0.23)$ | $0.74 (0.31)^*$ |
| INST | $-0.85 (0.40)^*$ | $-1.10 (0.55)^*$ |
| urban | $0.01 (0.01)$ | $-0.00 (0.01)$ |
| log(GDP) | $1.48 (0.56)^{**}$ | $2.48 (0.74)^{***}$ |
| log(pop) | $0.86 (0.15)^{***}$ | $2.30 (0.19)^{***}$ |
| Gov Eff | $-0.18 (0.17)$ | $-0.39 (0.23)^\dagger$ |
| Age 65+ | $0.05 (0.03)^\dagger$ | $0.07 (0.04)^\dagger$ |
| Dispersion $r$ | 1.49 | 0.84 |
| AIC | 651 | 1236 |
| Observations | 87 | 87 |

Two other cultural factors appear to affect COVID-19 death rates. Countries that value cosmopolitanism (COS) had more deaths from COVID-19 in the later stages of the outbreak (Table 1). This openness of national cultures to movement and visitors presumably helped spread COVID-19 [2, 18]. The cultural value of secular-rationalism (RAT) predicted fewer deaths in the first 10 days of the pandemic (Table 1), but had lost this effect after two weeks (Figure S3). We could speculate that cultural faith in science may have enabled national governments to to take early preventative action [2].

In the first two months of the outbreak, higher GDP per capita predicted higher COVID-19 death rates (Table 1, Figure S2). Although we find evidence, using new data [2], that higher GDP predicted a stronger and earlier government response to COVID-19 (Table S3), the weakness of government efficiency in our main regression (Table 1) suggests that such governments responses were not so effective in reducing the death rate. Instead, the reason GDP predicts more deaths may reflect economic incentives of wealthier populations to resist shutdown measures [22].

Among demographic controls, population size had the largest effect on COVID-19 deaths (Table 1). Populations with higher proportions of people over age 65 tended to have more deaths per capita. Percentage urban population did not have a significant effect when regressed with the other covariates (Table 1).

Finally, a larger question is the resilience of cultures and democracies to unprecedented challenges and ‘black swan’ events [11]. While trust in institutions predicted fewer COVID-19 deaths and ought to facilitate government action, this value has been declining for decades in many countries [13]. Cultural values of cosmopolitanism and openness, which underlie the economic prosperity and democracy of nations in the long term [21, 13, 14, 15] may in short-term crisis events hinder a strategic, coordinated national response. Hence, while a multi-decade trend towards greater openness towards minorities around the world [14] is encouraging, governments should consider the role of cultural values in preparing for the next pandemic.

## Data Availability

Original cultural values data generated for this research work have been archived within the Zenodo repository at the link below. Additional covariate data analysed during the current study are available in repositories hosted by (links below): World Bank repository, Our World in Data repositories for corronavirus statistics, Our World in Data repositories for urbanization, and the World Economic Forum Government Efficiency and Global Competitiveness Indices

https://doi.org/10.5281/zenodo.3559789

https://data.worldbank.org/data-catalog/world-development-indicators

https://ourworldindata.org/coronavirus

https://ourworldindata.org/urbanization

http://reports.weforum.org/global-competitiveness-index-2017-2018

## Acknowledgments

This research is supported by NSF grant 2028710. DJR was supported by a grant from the College of Arts and Sciences, University of Tennessee.

## Data availability

Original cultural values data [14] generated for this research work have been archived within the Zenodo repository: https://doi.org/10.5281/zenodo.3559789. Additional covariate data analysed during the current study are available in these repositories:

- World Bank repository, https://data.worldbank.org/data-catalog/world-development-indicators
- Our World in Data repositories for corronavirus statitstics, https://ourworldindata.org/coronavirus
- Our World in Data repositories for urbanization, https://ourworldindata.org/urbanization,
- World Economic Forum Government Efficiency and Global Competitiveness Indices http://reports.weforum.org/global-competitiveness-index-2017-2018.

## Supplementary Materials

### Cultural values predict national COVID-19 death rates

D.J. Ruck, J. Borycz, R.A. Bentley

## Materials and Methods

### Data

**Control variables** were collated by ourworldindata.com [27]. These variables included **percentage aged over 65 years** from the World Bank’s World Development Indicators, **population sizes** of nations in 2010 are from the United Nations Department of Economic and Social Affairs. **Percentage urban population** for nations in 2017 come from the World Bank’s development indicators [**?**].

**COVID-19 deaths** data from the European Centre for Disease Prevention and Control [27] were obtained from ourworldindata.com. **Government efficiency index** is taken from the World Economic Forum’s 2018 Global Competitiveness [26]; it is a composite measure that quantifies: 1) efficient public spending, 2) weak burdens on private companies, 3) efficient judiciary, 4) responsive to private sector and 5) transparent policy changes. **GDP per capita** is measured at at purchasing power parity (constant 2011 international dollars) for most recent year available for each country [28]. **Government response** index is a composite variable of comprising information on 17 policies thought to help mitigate COVID-19 spread [2]. Eight of the policies relate to containment (school closures, mobility restrictions etc); four relate to economic policies (direct payments etc) and five relate to heath policies (testing regimes, extra healthcare spending etc).

**Cultural values**, including secular-humanism (RAT), openness to minorities (COS) and trust in institutions (INST), were derived from multivariate statistics and the World and European Values surveys (WEVS) data from 109 nations [19, 20, 21, 13, 14, 15]. The WEVS data are derived from the same 64 questions in the five waves of these surveys at 5-year intervals since 1990, administered to 476,583 participants from 109 different nations. These data were compressed into multivariate factors in two steps. The first used Exploratory Factor Analysis (EFA) to identify nine cultural factors underlying the WEVS data. From the EFA step, we summarized the common variance in the WEVS data and thereby remove the portion of the total variance that is likely to be measurement error or other forms of statistical noise. The second step was to use the EFA factor loadings as weights for Principal Component Analysis (PCA) to derive principal components (PCs) that are orthogonal, which is advantageous for our subsequent regression models.

We used the first three of these cultural components in in our analysis here: ‘Trust in Institutions’ *INST*, ‘Openness to Minorities’ *COS* and ‘Secular-Humanism’ *RAT* [15]. These components were interpreted based on the correlated cultural factors from the raw survey questions. ‘Trust in Institutions’ *INST*, was correlated with cultural factors such as confidence in institutions *r* = 0.58 and interest in politics *r* = 0.86. This means that individuals with high trust in institutions report high confidence in institutions like the media, the army and government and also have an active interest in politics. The component we label Secular-Humanism, *RAT*, is correlated with secularism (*r* = 0.76), political engagement (*r* = 0.62), respect for individual rights (*r* = 0.59) and low prosociality (*r* = 0.45) [13]. This means that secular-humanist respondents to the WEVS are those who reported, for example, that religion is not important in their lives, that they are likely to attend protests or sign petitions, they only pay taxes when coerced and believe that homosexuality and divorce are justifiable [21, 13]. Openness to minorities, *COS*, is correlated with the exploratory cultural factors for ‘openness to out-groups’ (*r* = 0.78), ‘openness to norm violators’ (*r* = 0.78) and ‘subjective wellbeing’ (*r* = 0.43). High *COS* implies willingness to have neighbours that are immigrants, from another race, homosexual or from other stigmatized groups; as well as self-reporting a high level of happiness and satisfaction with life [13, 15].

## PCA and Multi-variate regressions

**Figure S1:**
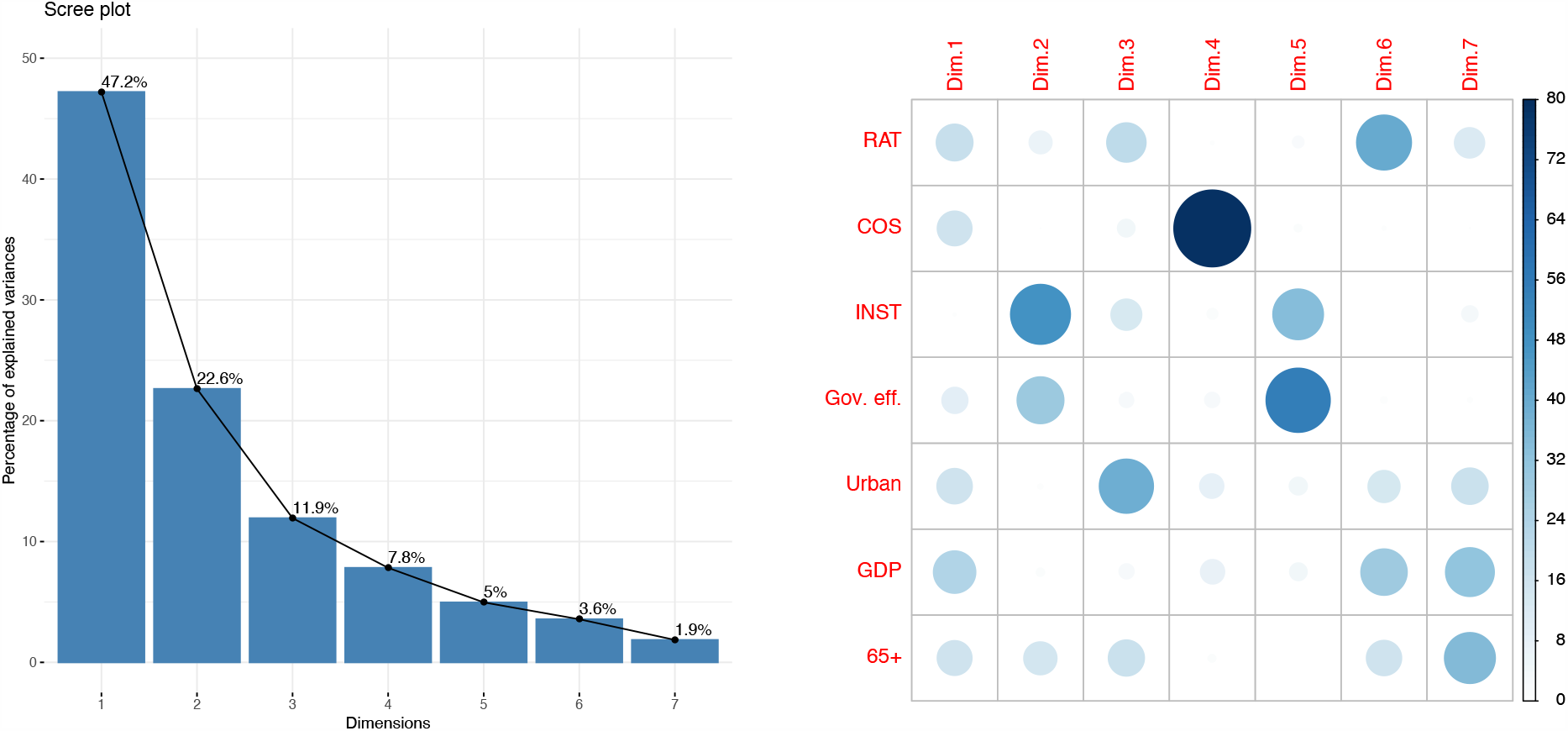
Left: Scree plot for the PCA analysis discussed in the text. Right: Bubble grid showing the relative loadings on the PCA.

**Figure S2:**
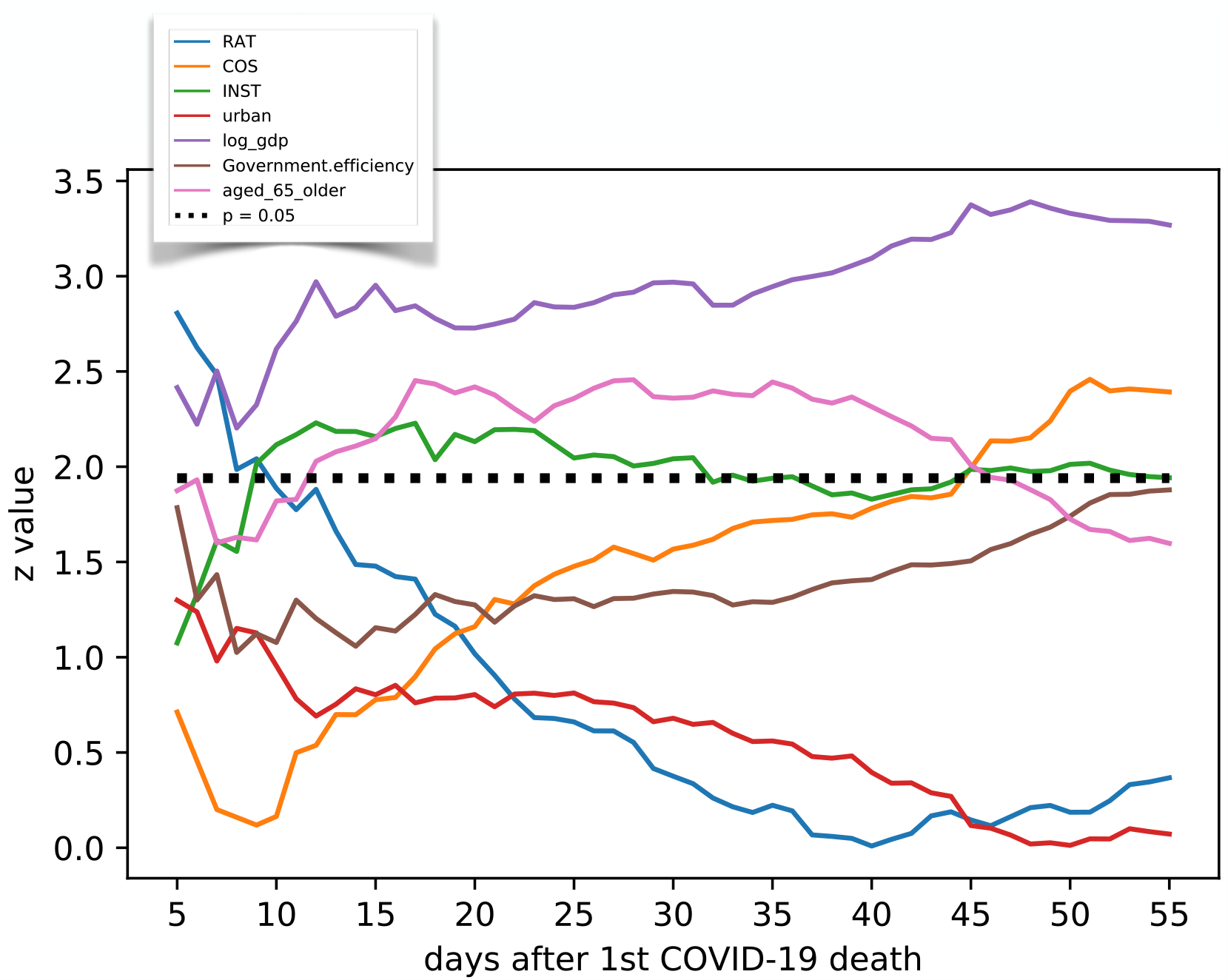
Significance of covariate effects on number of cases as the COVID-19 outbreak progresses; where z value = effect size/standard error. Dotted line indicates the z score corresponding with a p value of 0.05 in a two sided test.

For principal component analysis (PCA), we use the ‘Factominer’ and ‘Factoextra’ packages in R to compute the contributions (Table S1) and loadings (Table S2) of the principal components. The PCA included all variables for 83 nations excluding Kosovo, Serbia and Montenegro, North Ireland, and Taiwan, which lacked urbanization data.

**Table S1:** Variance explained by principal components and cumulative variance explained.

| Princ Comp: | PC 1 | PC 2 | PC 3 | PC 4 | PC 5 | PC 6 | PC 7 |
| --- | --- | --- | --- | --- | --- | --- | --- |
| Variance | 3.30 | 1.59 | 0.84 | 0.55 | 0.35 | 0.25 | 0.13 |
| % Variance | 47.2 | 22.6 | 11.9 | 7.8 | 5.0 | 3.6 | 1.9 |

**Table S2:** Loadings of the first three principal components (PC) on the variables. GDP is logarithm of GDP per capita; Age 65+ is fraction of population aged 65 and older.

| Variable | PC1 | PC2 | PC3 | PC4 |
| --- | --- | --- | --- | --- |
| RAT | 0.770 | -0.333 | 0.415 | -0.030 |
| COS | 0.728 | 0.010 | -0.189 | 0.656 |
| INST | 0.062 | 0.870 | 0.329 | 0.092 |
| Gov. Eff. | 0.552 | 0.680 | 0.154 | -0.128 |
| Urban | 0.742 | 0.083 | -0.571 | -0.210 |
| GDP | 0.889 | 0.123 | -0.159 | -0.206 |
| Age 65+ | 0.729 | -0.482 | 0.381 | -0.070 |

**Table S3:**
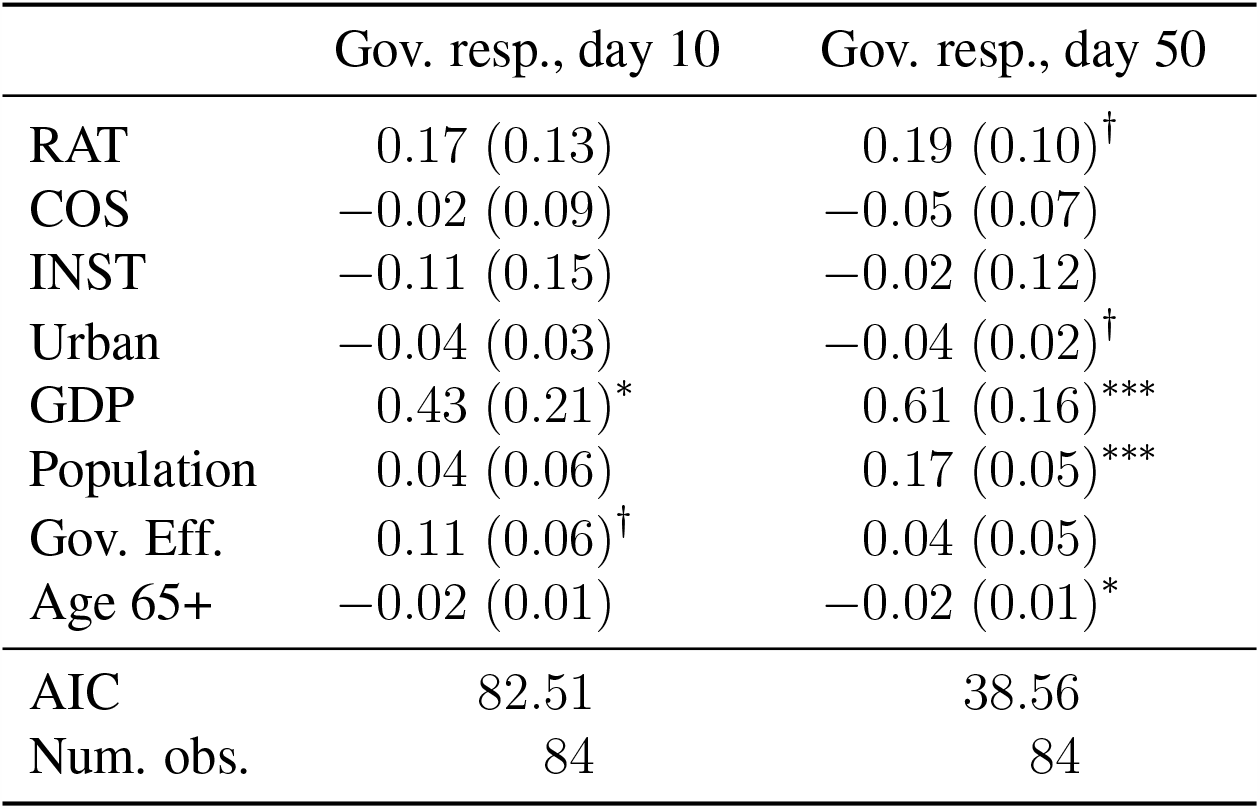
Results of multiple regression 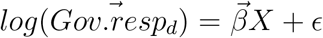, where *ϵ* ∼ *Normal*(0, *σ*) and covariate matrix *X* contains the covariates from the rows in this table. The vector 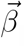 are the effects of covaraites on government response, both 10 and 50 days after outbreak. In parentheses are heteroskedastic standard errors. Heteroskedastic adjusted significance: ^***^*p*< 0.001, ^**^*p*< 0.01, ^*^*p*< 0.05, ^*†*^*p*< 0.10

|  | Gov. resp., day 10 | Gov. resp., day 50 |
| --- | --- | --- |
| RAT | 0.17 (0.13) | 0.19 (0.10)$^\dagger$ |
| COS | -0.02 (0.09) | -0.05 (0.07) |
| INST | -0.11 (0.15) | -0.02 (0.12) |
| Urban | -0.04 (0.03) | -0.04 (0.02)$^\dagger$ |
| GDP | 0.43 (0.21)* | 0.61 (0.16)*** |
| Population | 0.04 (0.06) | 0.17 (0.05)*** |
| Gov. Eff. | 0.11 (0.06)$^\dagger$ | 0.04 (0.05) |
| Age 65+ | -0.02 (0.01) | -0.02 (0.01)* |
| AIC | 82.51 | 38.56 |
| Num. obs. | 84 | 84 |

**Figure S3:**
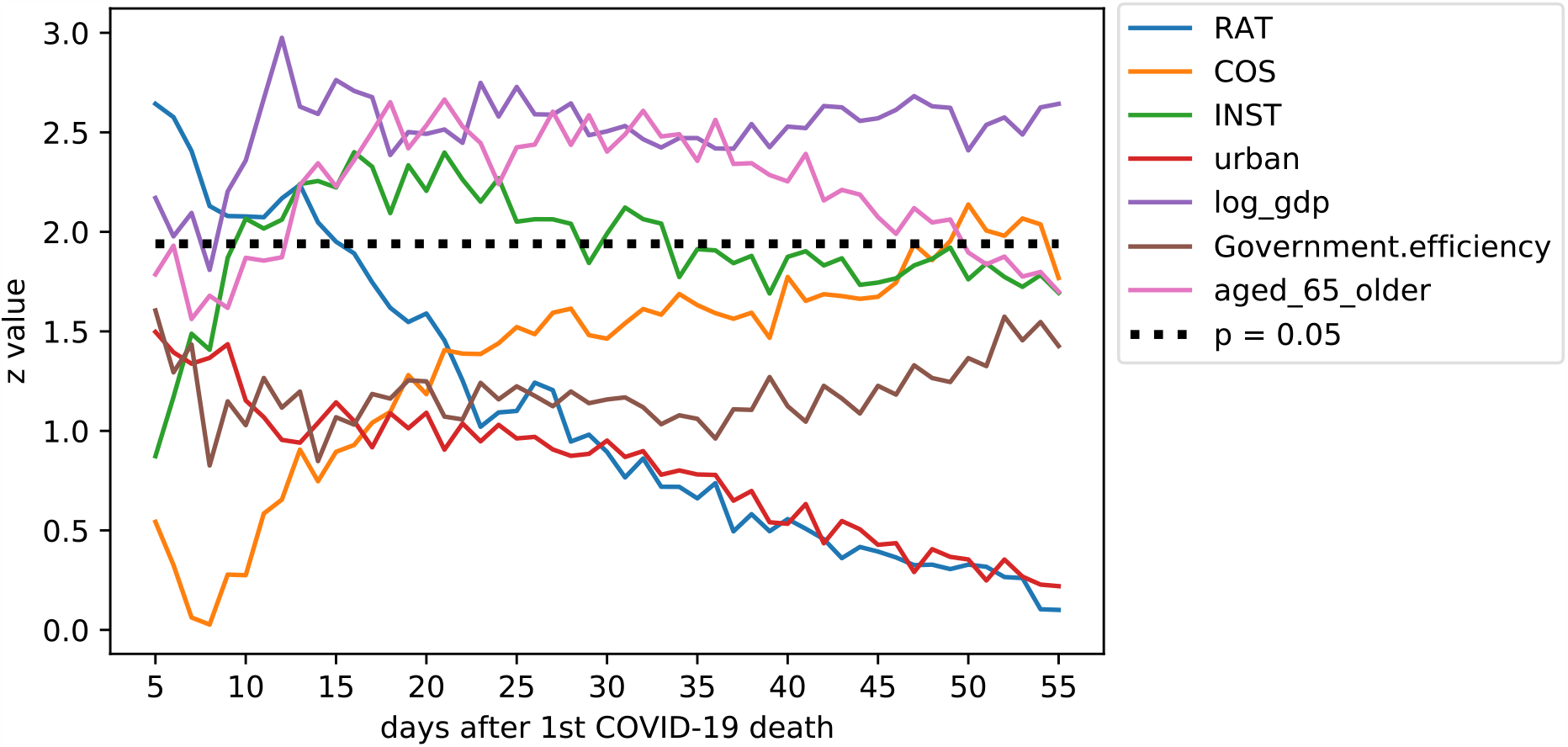
Significance of covariate effects using an ensemble of bootstrapped samples for parameter estimates; where z value = effect size/standard error on number of COVID-19 deaths. Dotted line indicates the z score corresponding with a p value of 0.05 in a two sided test.

**Table S4:**
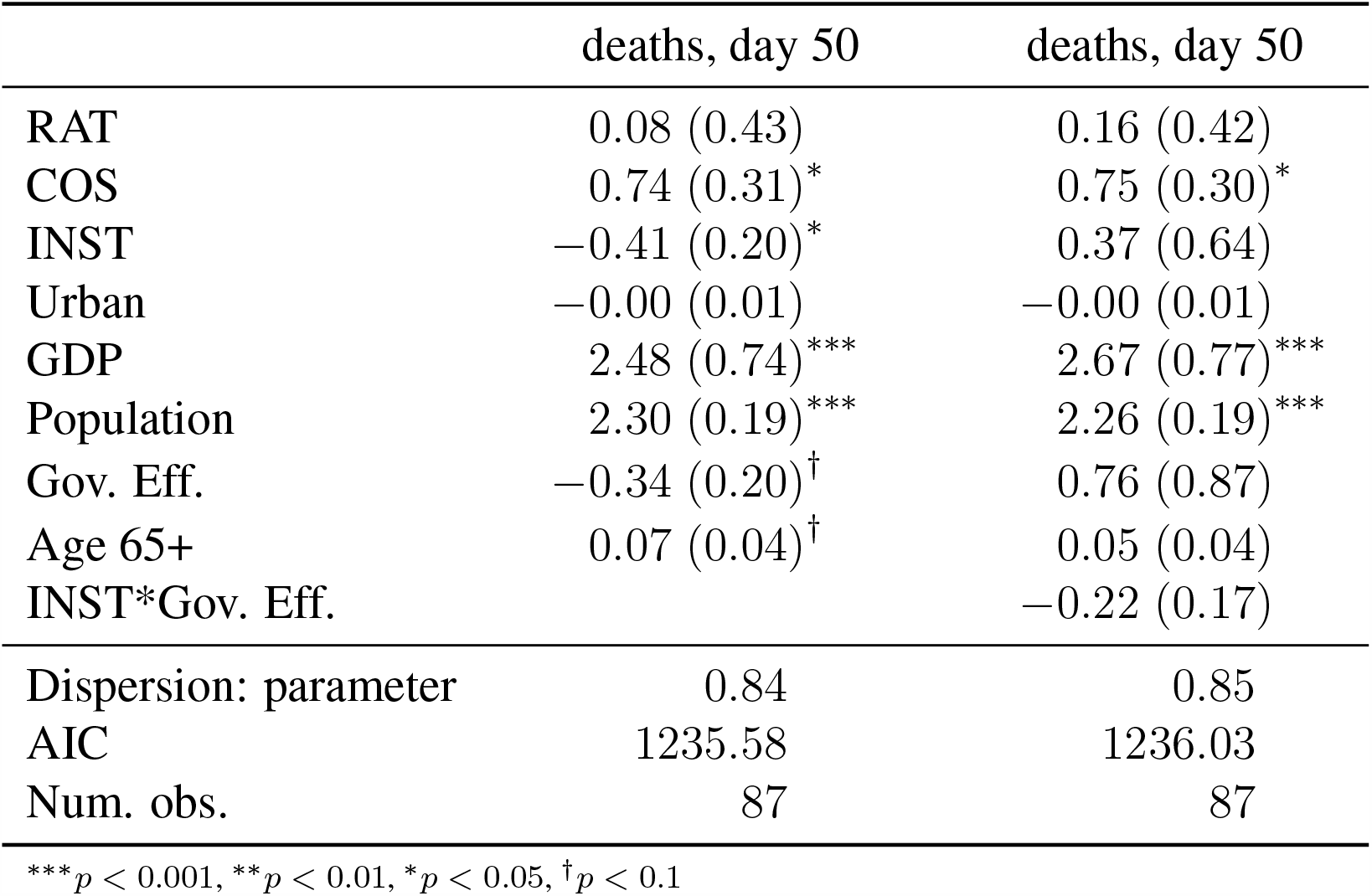
Negative binomial regression, including interaction between government efficiency and instiutional confidence. Predictors of COVID-19 deaths, 50 days after outbreak. AIC is not improved by adding the interaction term.

|  | deaths, day 50 | deaths, day 50 |
| --- | --- | --- |
| RAT | 0.08 (0.43) | 0.16 (0.42) |
| COS | 0.74 (0.31)* | 0.75 (0.30)* |
| INST | -0.41 (0.20)* | 0.37 (0.64) |
| Urban | -0.00 (0.01) | -0.00 (0.01) |
| GDP | 2.48 (0.74)*** | 2.67 (0.77)*** |
| Population | 2.30 (0.19)*** | 2.26 (0.19)*** |
| Gov. Eff. | -0.34 (0.20)<sup>†</sup> | 0.76 (0.87) |
| Age 65+ | 0.07 (0.04)<sup>†</sup> | 0.05 (0.04) |
| INST*Gov. Eff. |  | -0.22 (0.17) |
| Dispersion: parameter | 0.84 | 0.85 |
| AIC | 1235.58 | 1236.03 |
| Num. obs. | 87 | 87 |

\*\*\* $p < 0.001$ , \*\* $p < 0.01$ , \* $p < 0.05$ , <sup>†</sup> $p < 0.1$

